# Immune Microenvironment Profiling of Normal Appearing Colorectal Mucosa Biopsied Over Repeat Patient Visits Reproduciably Separates Lynch Syndrome Patients Based on Their History of Colon Cancer

**DOI:** 10.1101/2023.03.03.23286594

**Authors:** Rhonda M. Brand, Beth Dudley, Eve Karloski, Ashley Zyhowski, Rebecca Raphael, Danielle Pitlor, E. Jeffrey Metter, Reet Pai, Kenneth Lee, Randall E. Brand, Shikhar Uttam

## Abstract

**Introduction:** Lynch syndrome (LS) is the most common hereditary cause of colorectal cancer (CRC), increasing lifetime risk of CRC by up to 70%. Despite this higher lifetime risk, disease penetrance in LS patients is highly variable and most LS patients undergoing CRC surveillance will not develop CRC. Therefore, biomarkers that can correctly and consistently predict CRC risk in LS patients are needed to both optimize LS patient surveillance and help identify better prevention strategies that reduce risk of CRC development in the subset of high-risk LS patients.

**Methods:** Normal-appearing colorectal tissue biopsies were obtained during repeat surveillance colonoscopies of LS patients with and without a history of CRC, healthy controls (HC), and patients with a history of sporadic CRC. Biopsies were cultured in an *ex-vivo* explant system and their supernatants were assayed via multiplexed ELISA to profile the local immune signaling microenvironment. High quality cytokine signatures were identified using *rx*COV fidelity metric. These signatures were used to perform biomarker selection by computing their selection probability based on penalized logistic regression.

**Results:** Our study demonstrated that cytokine based local immune microenvironment profiling was reproducible over repeat visits and sensitive to patient LS-status and CRC history. Furthermore, we identified sets of biomarkers whose differential expression was predictive of LS-status in patients when compared to sporadic CRC patients and in identifying those LS patients with or without a history of CRC. Enrichment analysis based on these biomarkers revealed an LS and CRC status dependent constitutive inflammatory state of the normal appearing colonic mucosa.

**Discussion:** This prospective pilot study demonstrated that immune profiling of normal appearing colonic mucosa discriminates LS patients with a prior history of CRC from those without it, as well as patients with a history of sporadic CRC from HC. Importantly, it suggests existence of immune signatures specific to LS-status and CRC history. We anticipate that our findings have the potential to assess CRC risk in individuals with LS and help in preemptively mitigating it by optimizing surveillance and identifying candidate prevention targets. Further studies are required to validate our findings in an independent cohort of LS patients over multiple visits.

## 1 Introduction

Lynch Syndrome (LS) is the most common hereditary cause of colorectal cancer (CRC) with a prevalence of approximately 0.1-0.4% individuals in the general population and is responsible for 1-4% of patients with CRC (1). It is an autosomal dominant disease caused by a germline pathogenic variant (PV) in one of the DNA mismatch repair (MMR) genes (*MLH1, MSH2, MSH6* or *PMS2*) or deletions in the *EPCAM* gene that leads to silencing of *MSH2* via promoter hypermethylation. LS is characterized by a very rapid transformation along the adenoma-carcinoma sequence that usually occurs in 1-3 years in contrast to the 10–15-year timeline for MMR proficient tumors. Depending on the MMR genes involved, the lifetime risk of LS patients developing CRC is reported to be as high as 60% without surveillance (2). Given this heightened risk, the National Comprehensive Cancer Network (NCCN) recommends regularly scheduled surveillance colonoscopy be performed more frequently beginning at earlier ages for individuals with LS (3). Most LS patients undergoing high quality colonoscopy surveillance, however, do not develop CRC. This dichotomy raises the potential of safely lengthening colonoscopy surveillance intervals in a subset of LS patients deemed to be at lower risk of developing CRC, if they can be accurately identified.

CRC in LS patients display a highly microsatellite instable (MSI-H) phenotype, which is associated with increased immune infiltration of the CRC tumor microenvironment. This increase is in response to immunogenic frameshift peptides generated by the defective MMR machinery (4, 5). Interestingly, a systematic review of CRC literature indicates that LS-associated CRC tumors have an increased immune response when compared to sporadic MSI-H CRC tumors, even at the premalignant stage (6). A potential reason for this could be that normal appearing colonic crypts from LS patients can exhibit MMR deficiency (7-9). The presence of MMR deficient crypts, however, was independent of an LS patient’s cancer history (10). Such observations have resulted in an increasing realization that the immune status of normal colorectal mucosa in LS needs to be better characterized and understood. Toward this goal, a recent seminal study compared immune infiltration in tumor-distant normal appearing colorectal mucosa of LS patients with and without CRC with that of sporadic MSI-H and microsatellite-stable (MSS) CRC patients (11). Interestingly, it found elevated T-cell infiltration in normal mucosa of cancer-free LS patients when compared with MSS CRC patients adding to the evidence that immunogenic frameshift peptides can induce an antitumor immune profile in the absence of tumor. It also identified altered immune profiles between normal mucosa of cancer-free LS carriers and LS CRC patients and raised the possibility that the immune profile of normal mucosa may be a risk-modifier in LS patients and could potentially help improve patient surveillance.

To expand this concept, we sought to determine whether LS impacts the constitutive inflammatory state of colorectal tissue, with the long-term goal of identifying biomarkers to incorporate into CRC prevention approaches. Specifically, we profiled the cytokine-based, local immune signaling microenvironment of normal appearing colorectal mucosa from LS and non-LS patients without active CRC, but with or without a history of CRC. To strengthen the significance of our findings, we examined the reproducibility of our results at different time points. The annual to biannual endoscopic surveillance that LS patients undergo provided a unique source of tissue to both identify biomarkers and confirm their reproducibility over time, thereby demonstrating their robustness as biomarkers to integrate into prevention strategies. We anticipate that such characterization of the local immune signaling in normal appearing colorectal tissue from these patient cohorts will help advance our understanding of the interplay between immune microenvironment and risk of neoplastic progression in LS patients and help guide development of novel CRC cancer prevention strategies and risk-assessment approaches.

## 2 Materials and Methods

### 2.1 Patient Cohorts

LS, healthy control (HC) and non-LS patients with a history of colorectal cancer (Sporadic-CRC) but no active disease were recruited during screening and surveillance colonoscopies through an IRB-approved study at the University of Pittsburgh, Pittsburgh, PA (STUDY20010017). The LS cohorts consisted of individuals with an identified germline pathogenic/likely pathogenic variant in *MLH1, MSH2, MSH6, PMS2*, or *EPCAM* and either a personal history of colorectal cancer (LS-CRC) or no personal history of colorectal cancer (LS-noCRC). HC had no history of cancer (aside from non-melanoma skin cancer) within the last 5 years. Sporadic-CRC had documentation available that proves their colon tumor was microsatellite stable or that mismatch repair deficiency was somatic (e.g., MLH1 promoter hypermethylation). Individuals with a pathogenic or likely pathogenic variant in a different cancer susceptibility gene were also excluded from serving as controls. Individuals with active GI hemorrhage, an immunocompromised state, cardiopulmonary/hemodynamic instability, prior proctectomy, or who were prescribed anticoagulants were not approached for participation given the risk clinically unnecessary biopsies pose to these individuals.

### 2.2 Endoscopic Tissue Collection

Patients undergoing a scheduled clinical colonoscopy were approached at least 24 hours before their planned procedure by a single experienced gastroenterologist (REB) to obtain informed consent. Rectal biopsies were obtained with jumbo forceps in the proximal rectum (10 to 15 cm beyond dentate line) during their routine colonoscopy, which was performed at University of Pittsburgh Medical Center, Shadyside Hospital, Pittsburgh, PA. Those patients undergoing a repeat surveillance colonoscopy for clinical purposes were reconsented to obtain permission for repeat rectal biopsies.

### 2.3 Multiplexed ELISA on explant cultures

Biopsies collected from participants were immediately placed into tubes containing 20 mL of tissue transport media (tRPMI) comprised of RPMI 1640, 7.5% heat inactivated fetal bovine serum (HI-FBS), and 1% antibiotic-antimycotic (AB/AM) (Thermo Fisher Scientific, Grand Island, New York) and transported on ice until processed in the laboratory. Two biopsies from each participant were then immediately weighed and placed into individual wells of a 24-well culture plate, each containing 1 mL of complete media (cRPMI; RPMI containing 10% HI-FBS and 1% AB/AM) and incubated at 37°C with 5% CO_2._ Supernatants were collected after 24 hours, aliquoted and frozen at ≤-70°C as described earlier (12, 13).

Soluble biomarkers released from the explants into the supernatant through 24 h of culture were aliquoted and measured using mELISA (V-PLEXhuman biomarker 30-plex assay kit; Meso Scale Discovery, Rockville, MD). Measured biomarkers included Eotaxin, MIP-1β, Eoxtaxin-3, TARC, IP-10, MIP-1α, IL-8.1, MCP-1, MDC, MCP-4,GM-CSF, IL-1α, IL-5, IL-7, IL-12/IL-23p40, IL-15, IL-16, IL-17α, TNF-β, VEGF and proinflammatory IFN-γ, IL-1β, IL-2, IL-4, IL-6, IL-8.2, IL-10, IL-12p70, IL-13, TNF-α proteins commonly used to profile the immune signaling microenvironment. The presences two different IL-8s (IL-8.1 and IL8.2) reflect low and high dynamic ranges, respectively. All assays were performed according to the manufacturer’s instructions. To remove variations due to size of tissue biopsy, we normalized the assayed expression by tissue weight.

### 2.4 Computational Method

*Establishing biomarker fidelity*. As demonstrated in Figure 1, we applied *ratio-of-cross* coefficient of variation (*rx*COV) metric to establish the quality of the biomarker measurements. *rx*COV uses an objective threshold of zero to verify whether the good performance of a biomarker in separating between patient groups was not an assay associated experimental artifact (14). *rx*COV metric was independently applied to mELISA data from each visit.

**Figure 1.**
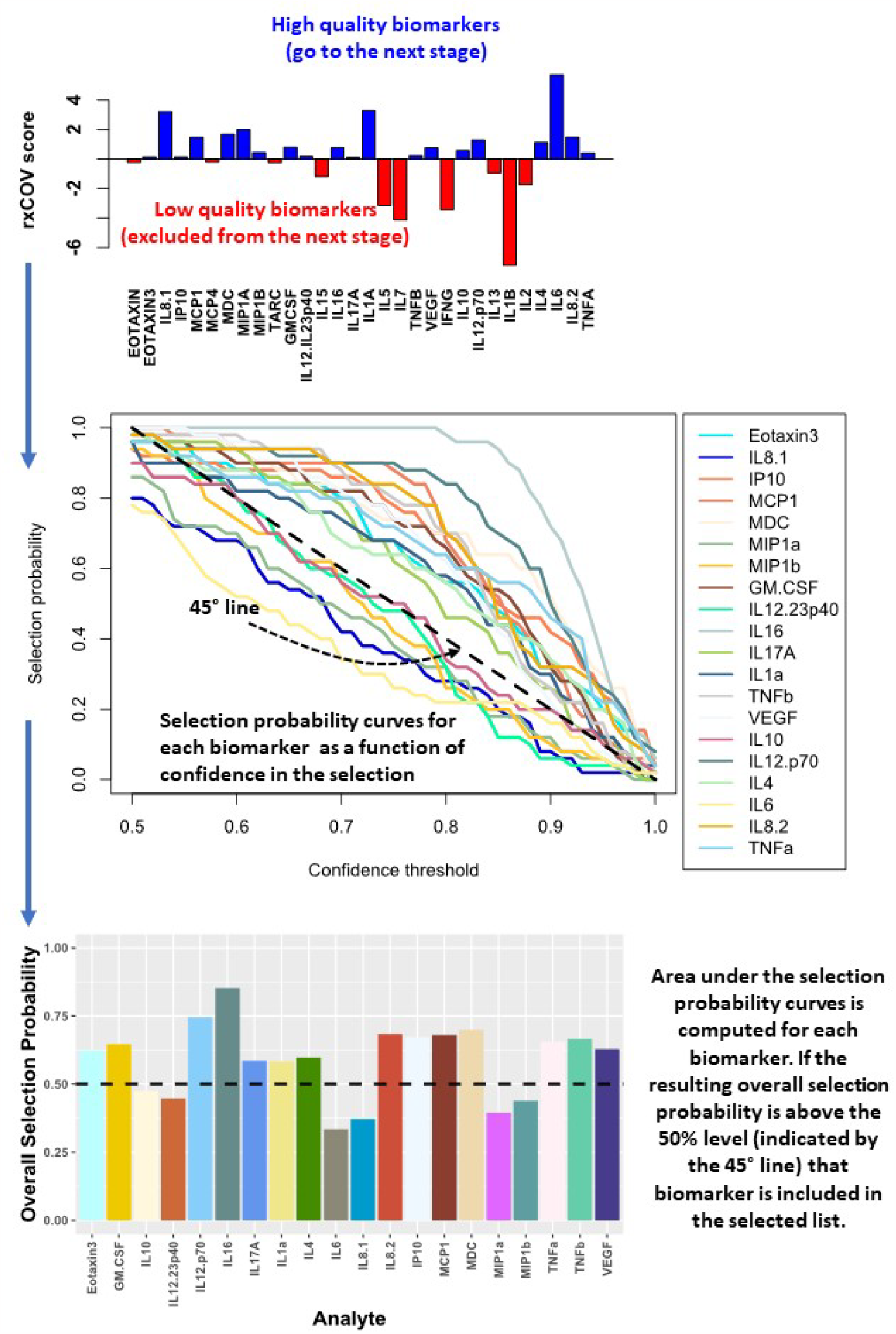
Computational method for biomarker selection. High fidelity biomarkers that satisfy the objective threshold of zero (shown in blue) are identified using the *rx*COV metric. Selection probability of these biomarkers is estimated using logistic regression with an elastic net penalty. The selection probability captures the ability of each biomarker in helping differentiate the patient groups being compared as a function of the selection confidence threshold. The area under the resulting curve is used to compute the overall selection probability for each biomarker. Those with overall selection probability greater than 0.5 comprise the selected biomarkers.

*Biomarker selection*. Biomarkers that passed the fidelity check were used to select a subset capable of separating the two patient groups being compared. The selection was performed using logistic regression with an elastic-net penalty (15). It was implemented using the glmnet R package (16, 17). Elastic-net penalty overcomes the shortcoming of lasso-based biomarker selection, which can randomly select a biomarker from a set of highly correlated biomarkers, while ignoring the rest of the correlated group. Since cytokine based signaling includes both autocrine and paracrine components and is pleiotropic in nature, lasso-based biomarker selection can be particularly deleterious in this context (15, 18). Elastic-net penalty on the other hand, can both perform efficient biomarker selection, while also accounting for grouping effect of highly correlated biomarkers. As a result, it excludes trivial biomarkers but performs grouped selection – selecting the whole group of correlated biomarkers, if some within the group are important in differentiating the two groups being compared. Such a selection process is better suited for cytokine-based biomarker selection. Specifically, we performed elastic-net-based penalized logistic regression utilizing two nested loops. The outer loop, with 100 iterations, sampled with replacement 70% of patients in each of the two groups being compared to generate a range of patient cohorts capturing the underlying patient distribution within each group. For each outer loop, an inner loop with 200 iterations was used to optimize the elastic-net penalized logistic regression model based on leave-one-out cross validation and estimate the selection probability for each biomarker based on its ability in consistently classifying the two patient groups. The final biomarker selection was performed by estimating their overall selection probabilities as a function of varying confidence thresholds. The advantage of this selection process is that it eschews use of an arbitrary threshold for biomarker selection and instead generates an overall selection-probability estimate of their ability to help differentiate the two patient groups. Biomarkers with more that 50% value on this scale were selected. Biomarker selection was independently performed for each visit.

*Prediction*: Random Forest (19) classifier was trained on 70% of the patient data and the remaining 30% was used for validation. Two hundred bootstraps were performed to capture the patient data distribution in the mutually exclusive training and validation set. The performance was measured using the area under the Receiver Operating Characteristic curve (aucROC). The classifier performance was independently computed for each visit.

## 3 Results

### 3.1 Patient demographics

The patient cohorts consisted of 15 HC, 14 Sporadic-CRC and 29 LS patients (16 LS-NoCRC and 13 LS-CRC). Demographic information including age, gender, variant and smoking status at the first visit are included in Table 1. No significant differences were observed between groups except that the Sporadic-CRC group was significantly older (p<0.017). As LS patients undergo regular screening colonoscopies, biopsies were collected from a second visit when possible (LS-CRC 13 of 13 patients; LS-NoCRC 12 of 16 patients), over a span of two colonoscopies separated by an average of 708±252 days. No repeat visits occurred for the HC and Sporadic-CRC patients.

**Table 1:** Demographics at First Biopsy

| **Group** | **HC**<br>**(n=15)** | **LS-NoCRC**<br>**(n=16)** | **LS-CRC**<br>**(n=13)** | **Sporadic-CRC**<br>**(n=14)** | **p value** |
| --- | --- | --- | --- | --- | --- |
| ***Gender*** |  |  |  |  | p=0.851 |
| Male | 9 | 4 | 4 | 5 |  |
| Female | 6 | 12 | 9 | 9 |  |
| ***Age at Biopsy*** | 51.3±14.5 | 50.6±14.5 | 59.6±7.6 | 63.9±12.8 | p=0.017* |
| ***BMI*** | 28.35 | 28.74 | 29.85 | 28.68 | p=0.902 |
| ***Pathogenic Variant*** |  |  |  |  | p=1.0 |
| MLH1 |  | 2 | 2 |  |  |
| MSH2 |  | 8 | 6 |  |  |
| MSH6 |  | 4 | 4 |  |  |
| PMS2 |  | 2 | 1 |  |  |
| ***Smoking Status*** |  |  |  |  | p=0.538 |
| Never Smoker | 6 | 9 | 9 | 9 |  |
| Former Smoker | 8 | 5 | 4 | 5 |  |
| Current Smoker | 1 | 2 | 0 | 0 |  |

### 3.2 Profiling the immune signaling microenvironment

To profile the impact of LS and CRC history on the constitutive inflammatory state of colorectal tissue, we directly compared the immune microenvironment in the following groups: **1)** LS patients with prior history of non-metastatic CRC (LS-CRC); **2)** LS patients with no prior history of malignancy (LS-NoCRC); Sporadic-CRC patients who had prior surgery for CRC (matched by type of resection to group 1); and Healthy controls (HC) defined as individuals with no personal history of hereditary syndrome or colorectal cancer who were undergoing colonoscopy for an indication of colorectal cancer screen and advanced adenoma, which was not identified. Using pairwise comparisons we sought to identify the impact of (a) LS on the microenvironment (group 2 vs 4), (b) LS on the microenvironment with a history of CRC in the background (group 1 vs 3), and (c) CRC history on the microenvironment in the presence of LS (group 1 vs 2). Importantly, we sought to determine the consistency of this impact over repeat patient visits spread over two years.

### 3.3 Multiplexed ELISA of *ex vivo* colorectal explant culture reproducibly profiles the local immune signaling microenvironment of colonic mucosa over repeat patient visits

We first established the reproducibility of our *ex-vivo* colorectal explant culture and multiplexed ELISA based profiling of the local immune signaling microenvironment in the colonic mucosa of LS patients over repeat visits. Toward this goal, we leveraged our ability to follow LS patients that are under annual to biannual endoscopic surveillance at our medical center. We obtained biopsies from those LS patients that did not experience any change in health status or develop any disease between their repeat visits spread over a period of two years (Figure 2A). The biopsies were cultured *ex-vivo* (see Material and Methods) and profiled using multiplexed ELISA. We next tested the fidelity of each of the cytokines comprising the immune profile using our *rx*COV metric to identify the subset of high-fidelity cytokines. Using pairwise Wilcoxon signed-rank test (Graphpad Prism, v9.0, Boston MA) (20), we compared the expression levels of each high-fidelity cytokine across the two visits. Our results show that over the nearly two-year period the expression levels of most cytokines remained stable with difference in expression only being statistically significant at the 0.05 level for 9/32 of the selected biomarkers (Figure 2B). Although, we are further expanding our temporal study to include additional visits, our initial findings indicate that we can reproducibly capture the constitutive inflammatory state of the local immune signaling microenvironment across repeat patient visits.

**Figure 2.**
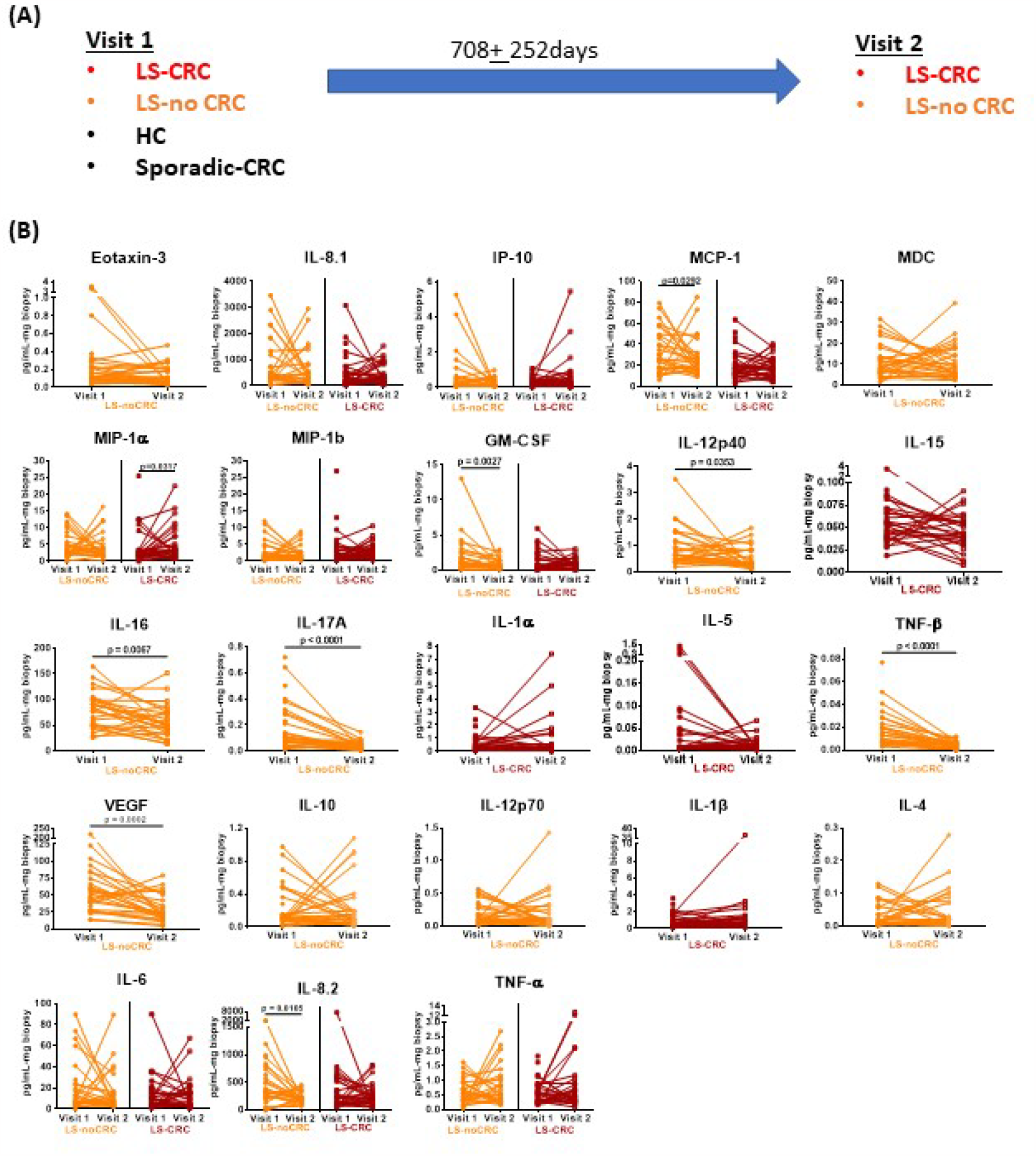
Reproducibility of cytokine profiles of the local immune signaling microenvironment over repeat patient visits. (**A**) Sketch of the repeat visits of LS patients under endoscopic surveillance. Biopsies obtained during these visits were profiled using *ex vivo* explant system and multiplexed ELISA. Biopsies from HC controls and non-LS patients with a history of CRC (sporadic-CRC patients) were obtained during a single visit (**B**) Pairwise comparison of biomarker expression between the two visits. Only the biomarkers that passed the rxCOV fidelity threshold are shown.

### 3.4 Profiling the impact of LS on the immune signaling microenvironment of patients *without* a history of CRC

We explored Lynch status associated modification of local immune signaling microenvironment of normal appearing mucosa in the absence of history of CRC. Using our computational method, we compared the cytokine profiles of LS patients without any history of CRC (LS-NoCRC) with the baseline healthy controls (HC) (Figure 3). We performed this comparison over two successive patient visits and identified Eotaxin-3, IL-16, IL-17A, IL-1α, and TNF-β as the set of cytokines that were consistently differentially modulated along the LS-status axis (Figure 3A). We tested the strength of this differential modulation by validating their ability to predict LS status of patients within the two cohorts utilizing Random Forest based classifier. Quantification of this performance via area under the Receiver Operating Characteristic curve (aucROC) revealed a relatively consistent level of performance over two patient visits that were separated by two years (Figure 3B). A direct comparison of expression levels of the six identified biomarkers from LS-NoCRC and HC are presented for visit 1 (Figure 3C) and LS-NoCRC visit 2 vs the single visit of HC (Figure 3D). We eschewed the typical statistical significance-based analysis that correlates differential signature with the outcome. Instead, we implemented an outcome driven biomarker selection strategy that selected those markers that were most capable of capturing the impact of LS without a history of CRC, while accounting for their pleiotropic and redundant activity. As a result, the differential expression of individual selected biomarkers captures the subtle effect of the impact of LS on the colorectal microenvironment even though it might not be statistically significant.

**Figure 3.**
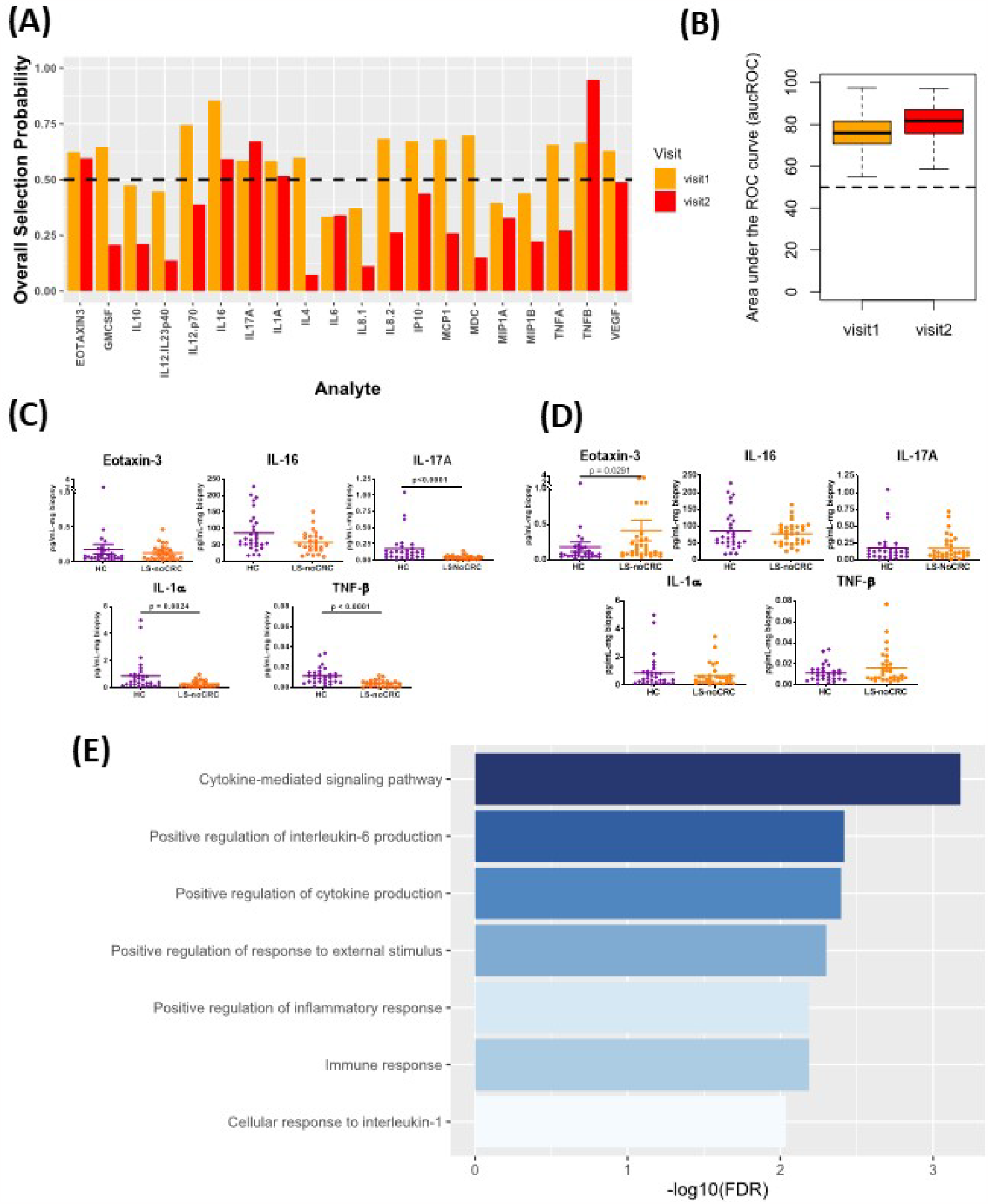
Cytokines capturing the impact of LS on the immune signaling microenvironment of patients without a history of CRC. (**A**) Biomarkers selected by our computational method for each visit. (**B**) Boxplots of area under the Receiver Operating Characteristic curve (aucROC) for the two visits. The boxplots were obtained using a Random Forest classifier trained on 70% of the patient data, with the remaining 30% used for validation. Two hundred bootstraps were performed to generate the boxplots and test the stability of the performance. Visit 1 aucROC: 75.6 (mean) ± 0.72 (se). Visit 2 aucROC: 80.7 (mean) ± 0.66 (se) (**C, D**) Expression levels of the selected cytokines for visit 1 and 2 respectively. (**E**) Gene ontology-based enrichment analysis.

Gene ontology-based enrichment analysis of the differential profile of Eotaxin-3, IL-16, IL-17A, IL-1α, and TNF-β revealed that, among other things, regulation of IL-6 production was fundamentally associated with LS status-based modification of the immune signaling microenvironment (21, 22)(Figure 3E). Interestingly, although IL-6 was not directly selected by our computational method, its enrichment suggests a potential close association between JAK/STAT signaling and LS status of the patient. Additionally, enrichment of IL-1 suggests the possibility of macrophage-mediated lymphocyte activation (23).

### 3.5 Profiling the impact of LS on the immune signaling microenvironment of patients *with* a history of CRC

We next explored modification of local immune signaling microenvironment of normal appearing colonic mucosa when the change in LS status was combined with a history of CRC. Specifically, we compared the cytokine profiles of Sporadic-CRC patient group with LS-CRC patients. Our computational analysis identified IP-10, MCP-1, MIP-1α, MIP-1β, IL-1α, IL-5, IL-1β, IL-6, IL-8.2 and TNFα as the set of biomarkers that consistently differentiated between the two groups over repeated visits using the same strategy detailed in the previous subsection (Figures 4A-D). Interestingly, presence of CRC background resulted in IL-6 being explicitly included in the selected list, suggesting its more explicit role in the normal colonic mucosa of patients with a CRC background.

**Figure 4.**
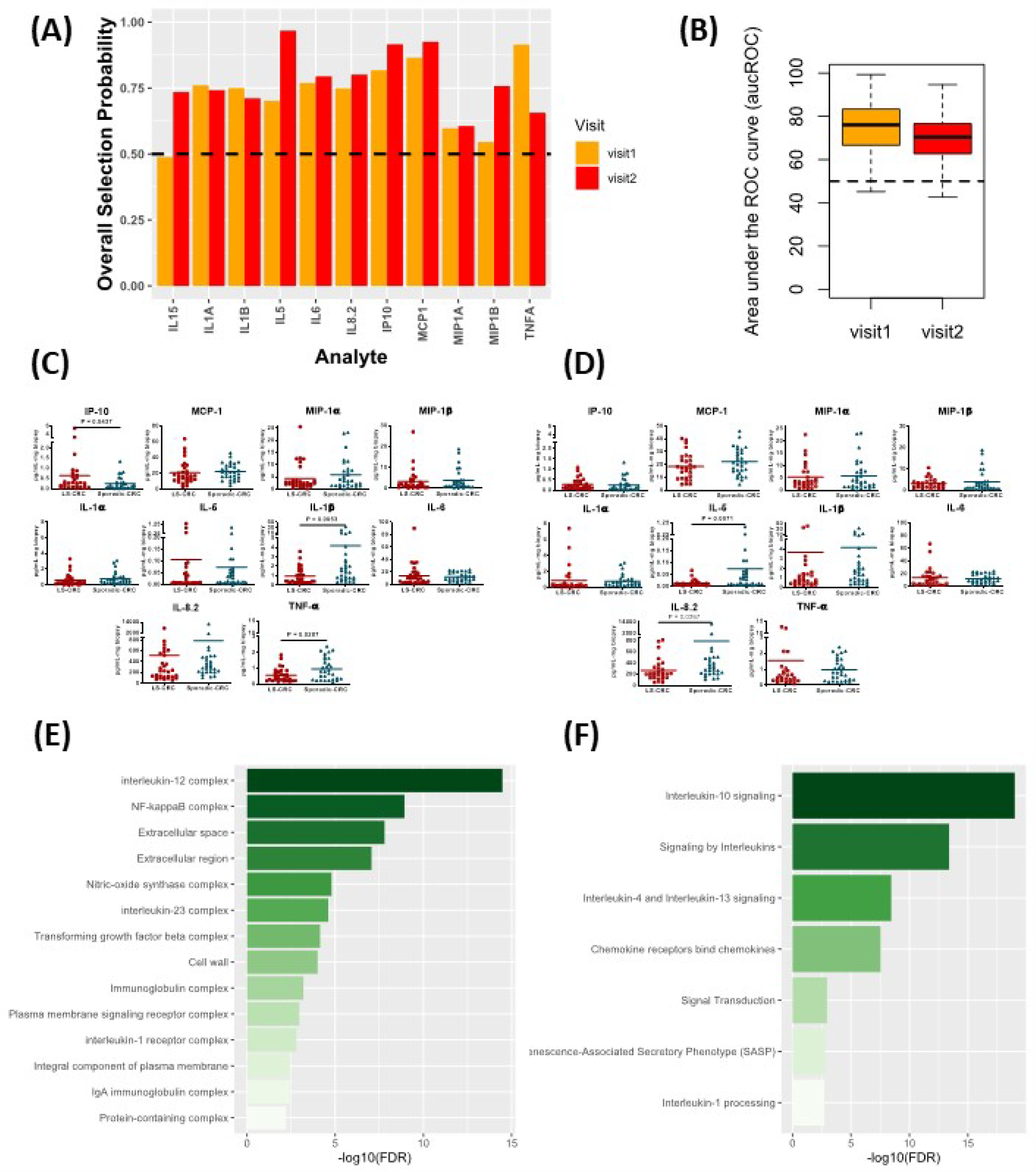
Cytokines capturing the impact of LS on the immune signaling microenvironment of patients with a history of CRC. (**A**) Biomarkers selected by our computational method for each visit. (**B**) Boxplots of area under the Receiver Operating Characteristic curve (aucROC) for the two visits. The boxplots were obtained using a Random Forest classifier trained on 70% of the patient data, with the remaining 30% used for validation. Two hundred bootstraps were performed to generate the boxplots and test the stability of the performance. Visit 1 aucROC: 74.3 (mean) ± 0.85 (se). Visit 2 aucROC: 69.3 (mean) ± 0.75 (se) (**C, D**) Expression levels of the selected cytokines for visit 1 and 2 respectively. (**E**) Gene ontology-based enrichment analysis. (**F**) Reactome pathway analysis.

Pathway analysis based on the selected biomarkers revealed an enriched role of IL-10, IL-4 and IL-13 signaling suggesting an immunosuppressive phenotype dependence (Figure 4E) (24). Concurrently, gene ontology-based enrichment analysis revealed an enriched role of IL-12, IL-23, NF-κB, and nitric oxide synthase (NOS) complexes that indicate a more proinflammatory phenotype (25, 26) and suggest a competition between the proinflammatory and immunosuppressive phenotypes in the normal colonic mucosa (Figure 4F). We note that many of these enriched complexes are implicated or dysregulated in patients with inflammatory bowel disease (27-30). Additionally, TGF-beta complex was also enriched. Aberration in TGF-beta signaling due mutations in TGF-beta receptors is commonly found in MMR deficient CRC, suggesting a potential residual effect of CRC history (31). Thus, our selected biomarkers seem to indicate that the impact of LS in the background of CRC is managed through a competition between the immunosuppressive and proinflammatory phenotypes along with residual effect of earlier CRC. The dominance of one phenotype over the other might potentially be concordant with increased risk of CRC relapse.

### 3.6 Profiling the impact of CRC history on immune signaling microenvironment of patients *with* LS

Finally, we studied the modification in the local immune signaling microenvironment of normal appearing colonic mucosa due to different CRC histories in patients with LS. Specifically, we profiled the LS-noCRC and LS-CRC patient groups. Our computational analysis identified IL-8.1, IP-10, MCP-1, GMCSF, and IL-1β as biomarkers that were consistently able to differentiate between the two groups in both visits (Figures 5A-D).

**Figure 5.**
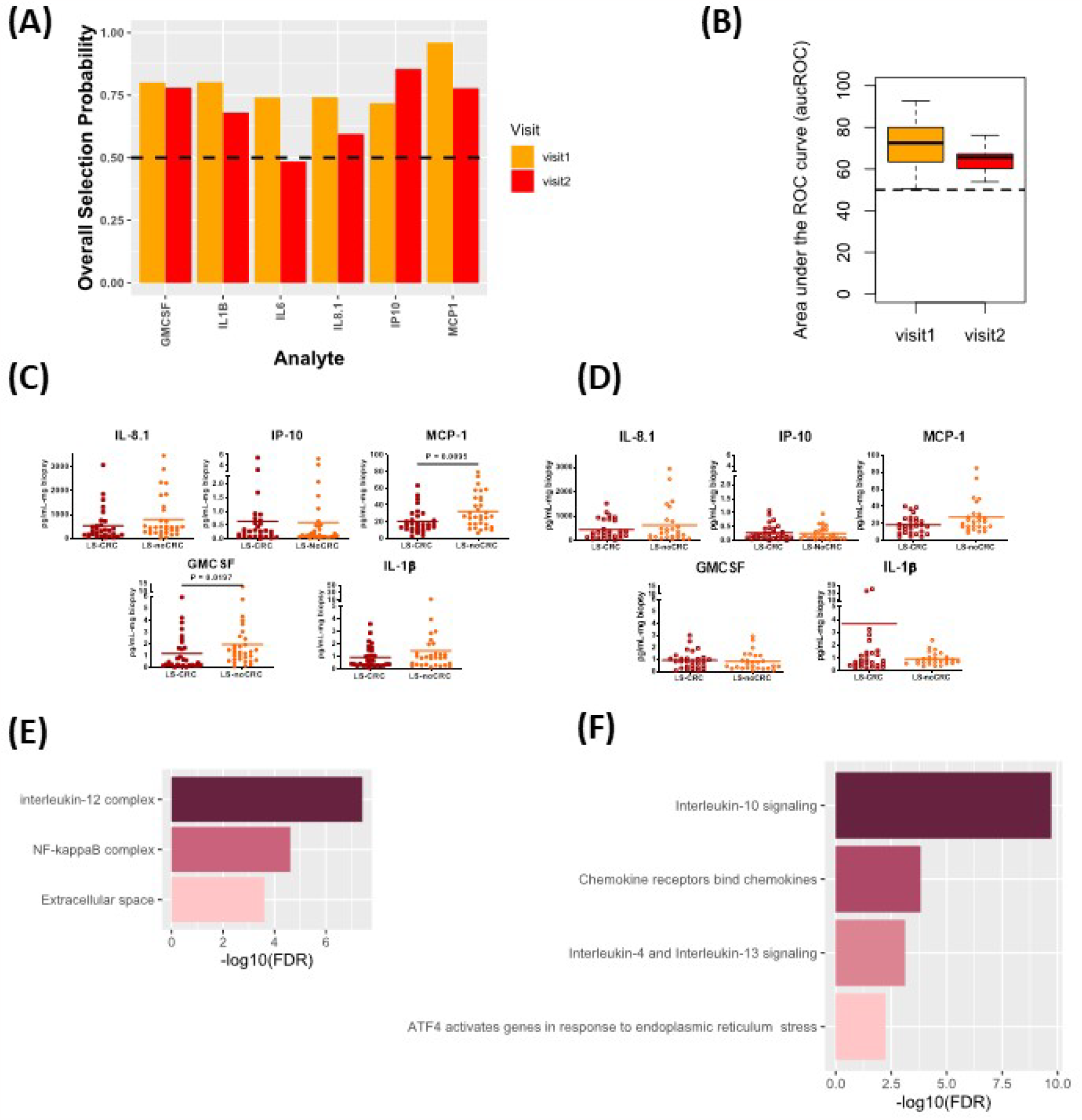
Cytokines capturing the impact of CRC history on immune signaling microenvironment of patients with LS. (**A**) Biomarkers selected by our computational method for each visit. (**B**) Boxplots of area under the Receiver Operating Characteristic curve (aucROC) for the two visits. The boxplots were obtained using a Random Forest classifier trained on 70% of the patient data, with the remaining 30% used for validation. Two hundred bootstraps were performed to generate the boxplots and test the stability of the performance. Visit 1 aucROC: 71.3 (mean) ± 0.9 (se). Visit 2 aucROC: 64.8 (mean) ± 0.48 (se) (**C, D**) Expression levels of the selected cytokines for visit 1 and 2 respectively. (**E**) Gene ontology-based enrichment analysis. (**F**) Reactome pathway analysis.

Pathway analysis combined with gene ontology-based enrichment analysis revealed enrichment of signaling pathways and molecular complexes that are a subset of those that capture the impact of differing LS status in patients with a history of CRC (Figures 5E, F). Direct comparison of Figure 4E with 5E, and of Figure 4F with 5F, seems to suggest that the impact on the local immune signaling microenvironment of normal appearing mucosa due to differing LS status in patients with CRC history is much broader than impact associated with differing CRC histories in LS patients. Importantly, we have now identified a consistent set of immune signaling signatures predictive of risk in both these settings over two patient visits. We aim to validate these signatures and better understand their biological underpinnings as part of our ongoing prospective study.

## 4 Discussion

Biomarkers that can correctly predict CRC risk in LS patients are needed to successfully mitigate the risk through prevention approaches including surveillance colonoscopy, vaccines, and chemoprevention. Ideally these approaches will be personalized with the goal of reducing costs by optimizing surveillance intervals or through predicting and monitoring agents that are most likely to prevent the development of colonic neoplasia. We took a novel approach to address this issue. Taking advantage of the large Lynch patient population that regularly undergoes surveillance colonoscopies at our institution, we were able to identify a cohort of patients that we followed over repeat visits. We obtained colorectal biopsies from these patients and using an *ex vivo* explant system combined with multiplexed ELISA, profiled the local immune signaling microenvironment of their normal appearing mucosa. Unlike most studies that focus on active disease in LS patients, our selection criteria mimicked two real-world surveillance scenarios for LS patients that are currently cancer-free: 1) those with no history of CRC and may develop CRC for the first time, or 2) those with a history of CRC that could experience a second primary CRC. By profiling the immune microenvironment of their normal appearing mucosa and using a history of CRC as a surrogate for CRC risk, our prospective and ongoing study aims to elucidate subtle but robust differences associated with immune modulation dependent on LS status and the residual effect of a prior resected CRC. Importantly our study established reproducibility of our results over repeat visits when there was no significant change in the patient’s health status, thereby, identifying a set of potential biomarkers that warrant further investigation in future prevention studies.

The potential of using differentially expressed serum and plasma cytokines as biomarkers for detecting the presence of CRC has been observed by multiple groups. By combining logistic regression models with multiplex ELISAs multiple teams have proposed a group of biomarkers capable of distinguishing between CRC and HC. Both the combination of serum levels of IL-9, Eotaxin, GM-CSF, and TNF-α and the combination of IL-4, IL-8, Eotaxin, IP-10, and TNF-α can distinguish between patients with CRC and HC (32). High serum IL-8, high IL-6, low MCP-1, low -IL1ra and low IP-10 were also able to distinguish between CRC patients with active disease from HC, with low serum IP-10 in combination with high IL-8 and IL-6 being associated with metastasized disease (33). However, these studies focus on detecting active disease, and not on characterizing alterations in the immune signaling microenvironment of normal mucosa in LS patients in a CRC-history dependent manner. By focusing on the latter, we anticipate that our findings have the potential to assess CRC risk and help in preemptively mitigating it by optimizing surveillance intervals and identifying immunomodulating prevention agents.

An important aspect of this study is that our analysis was informed by the redundant and pleiotropic nature of cytokine activity. We specifically avoided two pitfalls. First, we did not employ a simple p-value based determination of significance of differential cytokine expression to select important biomarkers. This is necessary due to the interconnectedness of cytokine signaling. It is possible that cytokines without a statistically significant differential expression between the comparison groups are important for separating those two groups. To avoid this pitfall, we utilized the predictive ability of the cytokines as the selection criterion. Second, while testing their predictive ability we utilized an elastic net penalty instead of the sometimes more commonly used lasso penalty to explicitly account for their correlated nature, while also excluding trivial associations.

Our analysis also benefited from our use of rxCOV metric, which we previously developed to avoid conflating assay-associated experimental variability with true significance of differential expression of any cytokine. Utilizing the rxCOV metric allowed us to filter out cytokines that truly were not statistically significant or whose differential expression was overwhelmed by assay associated noise. Although this latter aspect might have reduced the number of selected biomarkers for any two comparison groups, it did ensure that the differential expression of selected biomarkers truly had predictive ability in the context of our multiplexed ELISA measurements of explant colorectal mucosal cultures. The overall consistency of biomarkers selected between visit 1 and visit 2, which occurred nearly two years later, demonstrates the robustness of both the assay and analytical methods.

Due to the significant impact on healthcare resources, there is debate on the frequency that LS patients should undergo surveillance colonoscopy (34). Our long-term goal is to have a robust set of biomarkers that are strongly predictive of individual patient risk for developing colorectal neoplasia to identify those LS patients for whom surveillance intervals can be lengthened, based on their immune microenvironment detected from normal-appearing rectal biopsies. We note that although epidemiological studies have shown that risk of developing CRC in LS patients is correlated with MMR gene type, it remains unclear how to identify those patients in the genotype-defined cohorts that benefit from increased CRC surveillance. On the other hand, it is becoming increasingly probable that the local immune microenvironment of the normal colonic mucosa might be a more relevant, sensitive, and specific indicator of the evolving risk of a LS patient developing colorectal neoplasia (11). It is also potentially better indicator than serum biomarker levels that characterize systemic alterations that are not specific to the subtle alterations in the local microenvironment. Our approach of utilizing the patient biopsy obtained during regularly scheduled surveillance to profile their local immune microenvironment, therefore, has the potential to be incorporated into managing patient surveillance intervals.

Our study is currently limited by lack of complementary imaging data that corroborates cytokine activity differences in the context of immune cell infiltrates and their states of activation and polarization. Future work will focus on addressing this aspect while continuing to build on our strength of being able to follow patients over multiple visits, increasing the relatively small number of patients and expanding our cytokine repertoire. Combining these findings with sequencing-based analysis will help us not only optimize surveillance strategies but also identify potential immunoprevention candidate targets that will have the benefit of a long follow-up.

This study was focused on characterizing the local immune signaling microenvironment of normal appearing mucosa in clinically relevant LS patient groups and utilizing this characterization to identify a set of biomarkers that were consistently able to differentiate patients in an LS and CRC status dependent manner over two visits. The identification of our candidate biomarkers requires validation in a prospective, blinded studies using an independent cohort of LS patients. Further work is required to determine if the biomarkers can play a role in selecting prevention strategies such as identifying potential biological pathways that should be targets or monitoring response by detecting alterations in the immune signaling microenvironment.

## Data Availability

All data produced in the present study are available upon reasonable request to the authors.

## Acknowledgements

This project was supported in part by the Hillman Developmental Funds (P30CA047904). The authors would like to recognize the study participants and thank the clinical coordinators (Nancy Abubaker, Marietta Kocher, and Parker Ulrich) and laboratory staff (Katrina Culbertson, Alyssa Hein, Katie Sauka, Aaron Siegel, and Amanda Swistok) for their assistance in patient recruitment, sample collection and tissue assays.

## Author contributions

R.E.B, R.M.B, E.J.M, B.D., R.P, K.L. and S.U. conceived the concept; B.D., E.K. and R.E.B. collected the patient samples; R.M.B. and A.Z. performed the tissue assays; R.M.B, E.J.M, A.Z., R.R., D.P. and S.U. performed the analyses; R.E.B, R.M.B and S.U. wrote the manuscript. All authors reviewed the manuscript.

## References

1. Pastor DM, Schlom J. Immunology of Lynch Syndrome. Curr Oncol Rep. 2021;23(8):96.

2. Samadder NJ, Baffy N, Giridhar KV, Couch FJ, Riegert-Johnson D. Hereditary Cancer Syndromes-A Primer on Diagnosis and Management, Part 2: Gastrointestinal Cancer Syndromes. Mayo Clin Proc. 2019;94(6):1099–116.

3. Weiss JM, Gupta S, Burke CA, Axell L, Chen LM, Chung DC, et al. NCCN Guidelines(R) Insights: Genetic/Familial High-Risk Assessment: Colorectal, Version 1.2021. J Natl Compr Canc Netw. 2021;19(10):1122–32.

4. Schwitalle Y, Linnebacher M, Ripberger E, Gebert J, von Knebel Doeberitz M. Immunogenic peptides generated by frameshift mutations in DNA mismatch repair-deficient cancer cells. Cancer Immun. 2004;4:14.

5. Willis JA, Reyes-Uribe L, Chang K, Lipkin SM, Vilar E. Immune Activation in Mismatch Repair-Deficient Carcinogenesis: More Than Just Mutational Rate. Clin Cancer Res. 2020;26(1):11–7.

6. Bohaumilitzky L, von Knebel Doeberitz M, Kloor M, Ahadova A. Implications of Hereditary Origin on the Immune Phenotype of Mismatch Repair-Deficient Cancers: Systematic Literature Review. J Clin Med. 2020;9(6).

7. Pai RK, Dudley B, Karloski E, Brand RE, O’Callaghan N, Rosty C, et al. DNA mismatch repair protein deficient non-neoplastic colonic crypts: a novel indicator of Lynch syndrome. Mod Pathol. 2018;31(10):1608–18.

8. Staffa L, Echterdiek F, Nelius N, Benner A, Werft W, Lahrmann B, et al. Mismatch repair-deficient crypt foci in Lynch syndrome--molecular alterations and association with clinical parameters. PLoS One. 2015;10(3):e0121980.

9. Kloor M, Huth C, Voigt AY, Benner A, Schirmacher P, von Knebel Doeberitz M, et al. Prevalence of mismatch repair-deficient crypt foci in Lynch syndrome: a pathological study. Lancet Oncol. 2012;13(6):598–606.

10. Brand RE, Dudley B, Karloski E, Das R, Fuhrer K, Pai RK, et al. Detection of DNA mismatch repair deficient crypts in random colonoscopic biopsies identifies Lynch syndrome patients. Fam Cancer. 2020;19(2):169–75.

11. Bohaumilitzky L, Kluck K, Hüneburg R, Gallon R, Nattermann J, Kirchner M, et al. The Different Immune Profiles of Normal Colonic Mucosa in Cancer-Free Lynch Syndrome Carriers and Lynch Syndrome Colorectal Cancer Patients. Gastroenterology. 2022;162(3):907-19.e10.

12. Brand RM, Biswas N, Siegel A, Myerski A, Engstrom J, Jeffrey Metter E, et al. Immunological responsiveness of intestinal tissue explants and mucosal mononuclear cells to ex vivo stimulation. J Immunol Methods. 2018;463:39–46.

13. Brand RM, Siegel A, Myerski A, Metter EJ, Engstrom J, Brand RE, et al. Ranpirnase Reduces HIV-1 Infection and Associated Inflammatory Changes in a Human Colorectal Explant Model. AIDS Res Hum Retroviruses. 2018;34(10):838–48.

14. Brand RM, Pitlor D, Metter EJ, Dudley B, Karloski E, Zyhowski A, et al. rxCOV is a quantitative metric for assessing immunoassay analyte fidelity. Sci Rep. 2023;13(1):88.

15. Zou H, Hastie T. Regularization and variable selection via the elastic net. Journal of the Royal Statistical Society: Series B (Statistical Methodology). 2005;67(2):301–20.

16. Friedman J, Hastie T, Tibshirani R. Regularization Paths for Generalized Linear Models via Coordinate Descent. J Stat Softw. 2010;33(1):1–22.

17. Simon N, Friedman J, Hastie T, Tibshirani R. Regularization Paths for Cox’s Proportional Hazards Model via Coordinate Descent. J Stat Softw. 2011;39(5):1–13.

18. Jiang P, Zhang Y, Ru B, Yang Y, Vu T, Paul R, et al. Systematic investigation of cytokine signaling activity at the tissue and single-cell levels. Nat Methods. 2021;18(10):1181–91.

19. Breiman L. Random Forests. Machine Learning. 2001;45(1):5–32.

20. Pratt JW, Gibbons JD. Concepts of nonparametric theory. New York: Springer-Verlag; 1981. xvi, 462 p. p.

21. The Gene Ontology resource: enriching a GOld mine. Nucleic Acids Res. 2021;49(D1):D325–d34.

22. Ashburner M, Ball CA, Blake JA, Botstein D, Butler H, Cherry JM, et al. Gene ontology: tool for the unification of biology. The Gene Ontology Consortium. Nat Genet. 2000;25(1):25–9.

23. Kaneko N, Kurata M, Yamamoto T, Morikawa S, Masumoto J. The role of interleukin-1 in general pathology. Inflamm Regen. 2019;39:12.

24. Gillespie M, Jassal B, Stephan R, Milacic M, Rothfels K, Senff-Ribeiro A, et al. The reactome pathway knowledgebase 2022. Nucleic Acids Research. 2021;50(D1):D687–D92.

25. Baig MS, Zaichick SV, Mao M, de Abreu AL, Bakhshi FR, Hart PC, et al. NOS1-derived nitric oxide promotes NF-kappaB transcriptional activity through inhibition of suppressor of cytokine signaling-1. J Exp Med. 2015;212(10):1725–38.

26. Liu T, Zhang L, Joo D, Sun SC. NF-kappaB signaling in inflammation. Signal Transduct Target Ther. 2017;2:17023-.

27. Kolios G, Valatas V, Ward SG. Nitric oxide in inflammatory bowel disease: a universal messenger in an unsolved puzzle. Immunology. 2004;113(4):427–37.

28. Aggeletopoulou I, Assimakopoulos SF, Konstantakis C, Triantos C. Interleukin 12/interleukin 23 pathway: Biological basis and therapeutic effect in patients with Crohn’s disease. World J Gastroenterol. 2018;24(36):4093–103.

29. Eftychi C, Schwarzer R, Vlantis K, Wachsmuth L, Basic M, Wagle P, et al. Temporally Distinct Functions of the Cytokines IL-12 and IL-23 Drive Chronic Colon Inflammation in Response to Intestinal Barrier Impairment. Immunity. 2019;51(2):367–80 e4.

30. Atreya I, Atreya R, Neurath MF. NF-kappaB in inflammatory bowel disease. J Intern Med. 2008;263(6):591–6.

31. Itatani Y, Kawada K, Sakai Y. Transforming Growth Factor-beta Signaling Pathway in Colorectal Cancer and Its Tumor Microenvironment. Int J Mol Sci. 2019;20(23).

32. Yamaguchi M, Okamura S, Yamaji T, Iwasaki M, Tsugane S, Shetty V, et al. Plasma cytokine levels and the presence of colorectal cancer. PLoS One. 2019;14(3):e0213602.

33. Kantola T, Klintrup K, Väyrynen JP, Vornanen J, Bloigu R, Karhu T, et al. Stage-dependent alterations of the serum cytokine pattern in colorectal carcinoma. Br J Cancer. 2012;107(10):1729–36.

34. Engel C, Vasen HF, Seppala T, Aretz S, Bigirwamungu-Bargeman M, de Boer SY, et al. No Difference in Colorectal Cancer Incidence or Stage at Detection by Colonoscopy Among 3 Countries With Different Lynch Syndrome Surveillance Policies. Gastroenterology. 2018;155(5):1400-+.

